# Tuberculosis Infection Control in MDR-TB designated hospitals, Jiangsu Province of China

**DOI:** 10.1101/2024.09.11.24313499

**Authors:** Honghuan Song, Guoli Li, Zhuping Xu, Feixian Wang, Xiaoping Wang, Bing Dai, Xing Zhang, Jincheng Li, Yan Li, Limei Zhu

## Abstract

**Background:** Hospital-acquired TB (Tuberculosis) infection among healthcare workers (HCWs) and patients is a severe problem due to the increased attributable risk of TB infection among these groups.

**Methods:** A standardized tool was applied. The assessment was conducted by direct observation, document review, and interviews with the facility heads. Baseline evaluation of TBIC (Tuberculosis infection control) in TB outpatient, inpatient departments, and laboratories was completed by January 2019. Based on the results, we implemented a comprehensive package of interventions, including administrative, environmental engineering, and respiratory protection (PPE) three-level hierarchy of controls. Subsequent monitoring was finalized quarterly and improvement measures should be formulated accordingly. More than two years of follow-up data was collected until August 31, 2021, by hospitals, municipality CDCs, and Jiangsu provincial CDC.

**Results:** At baseline, the implementation rate of administrative, environmental engineering and PPE IC was 57.29%, 59.21%, and 66.63%, respectively. After evaluation and implementation, priority way for cough patients was established, mechanical ventilation and the use of masks were improved, UV and UVGI lights were settled in need. The implementation rate of administrative, environmental and PPE IC were significantly increased to 86.27%, 87.41%, and 98.42%(*P*<0.05), respectively.

**Conclusions:** After more than one and a half years of intervention, TBIC in the designated hospitals has significantly improved. However, the availability of separate TB wards remains suboptimal. TB IC measures must be strengthened to reduce TB transmission among HCWs and non-TB patients. This method was practical and suitable to be popularized in countries with high TB burden.

## Introduction

China still with high tuberculosis (TB) burden[1]. Similar to other countries with a high TB burden, hospital-acquired TB has long been recognized as an important infection hazard for health care workers (HCWs) and other non-TB patients, due to the increased attributable risk of TB infection among these groups[2-4]. Therefore, infection prevention and control (IPC) measures are needed, aim to reduce the hospital-acquired TB risk.

Jiangsu Province locates in eastern China and is known as an eastern-central coastal province of the People’s Republic of China. It is one of the leading provinces in finance, education, technology, and tourism. In 2020, the incidence rate of tuberculosis in Jiangsu is half of the national average level, and the number of tuberculosis cases was more than 20 thousand, which was 8.97% lower than in 2019. Each prefecture-level city from Jiangsu Province set up a designated diagnosis and treatment hospital for drug-resistant TB.

It is evidenced that healthcare workers (HCW) are at higher risk to acquire TB than the general population, due to their possible occupational exposure to TB patients[2]. Furthermore, TB transmission in hospitals not only contributes to the risk for HCW but also to fellow patients and visitors[5].

Although the World Health Organization (WHO) and China CDC have issued recommendations for TB infection control within health facilities, limited available resources preclude the implementation of more efficient controls, such as engineering controls and personal protective equipment.

For this indication, a comprehensive technical support package was introduced in 6 drug-resistant TB Designated Hospitals in Jiangsu Province of China, including Suzhou Fifth People’s Hospital, Wuxi Fifth People’s Hospital, Changzhou Third People’s Hospital, Yangzhou Third People’s Hospital, Zhenjiang Third People’s Hospital and Nantong Sixth People’s Hospital which locate in southern Jiangsu, Central Jiangsu and Northern Jiangsu, respectively. The present study aimed to assess the extent of implementation of TBIC measurements before and after the introduction of this package.

## Methods

### 1. Study design

The measurements used in this study were based on the TB comprehensive pilot project jointly carried out by Jiangsu CDC and China CDC. A set of standardized assessment tools were developed, according to the WHO guidelines on tuberculosis infection prevention and control (2019 update), and the WHO recommendations for implementation of TBIC in healthcare settings. These tools addressed various aspects of administrative controls, environmental controls as well as the use of respiratory protection (personal protective equipment, PPE),including 18, 16 and 6 independent test items, respectively. A before-after study to improve the status of TB infection control measures implementation was conducted from December 1 2019 to September 1, 2021, in the TB outpatient department, inpatient department, and laboratory of the six drug-resistant designated hospitals in Jiangsu province, China, they are: Suzhou Fifth People’s Hospital, Wuxi Fifth People’s Hospital, Changzhou Third People’s Hospital, Yangzhou Third People’s Hospital, Zhenjiang Third People’s Hospital and Nantong Sixth People’s Hospital.

All assessments were carried out by the same investigators. The key information was individually collected and recorded at all sites. Based on the findings of the baseline assessment, we implemented a comprehensive package of interventions, including administrative, environmental engineering, and respiratory protection (PPE) three-level hierarchy of controls. After that, monitoring was done quarterly, improvement measures should be formulated according to the monitoring results. After six quarterly assessments of follow-up, final assessment data was collected by the hospitals, Municipal CDC, and Jiangsu CDC, respectively. Due to the impact of the COVID-19 pandemic, time to complete the assessment varies by institution.

### 2. Data management and statistical analysis

Data was collected in Excel tables and converted into scores. Scores according to the on-site evaluation of each indicator, 2 points for completed, 1 point for in progress, and 0 for uncompleted. Implementation rate = SUM(scores) / (2*(count).The proportions were calculated using SPSS version 24.0 (IBM Corp., Armonk, New York, USA).

## Results

### 1. The improvement of TBIC in the designated hospitals

At baseline, the implementation rate of administrative, environmental engineering and PPE IC was 57.29%, 59.21% and 66.63%, respectively. After evaluation and implementation, priority way for cough patients was established, mechanical ventilation, and the use of masks was improved, UV lights were settled in need, UVGI were settled in the TB consulting rooms. The implementation rate of administrative, environmental and PPE IC were significantly increased to 86.27%, 87.41% and 98.42%, respectively (FIG 1).

**Fig 1.**
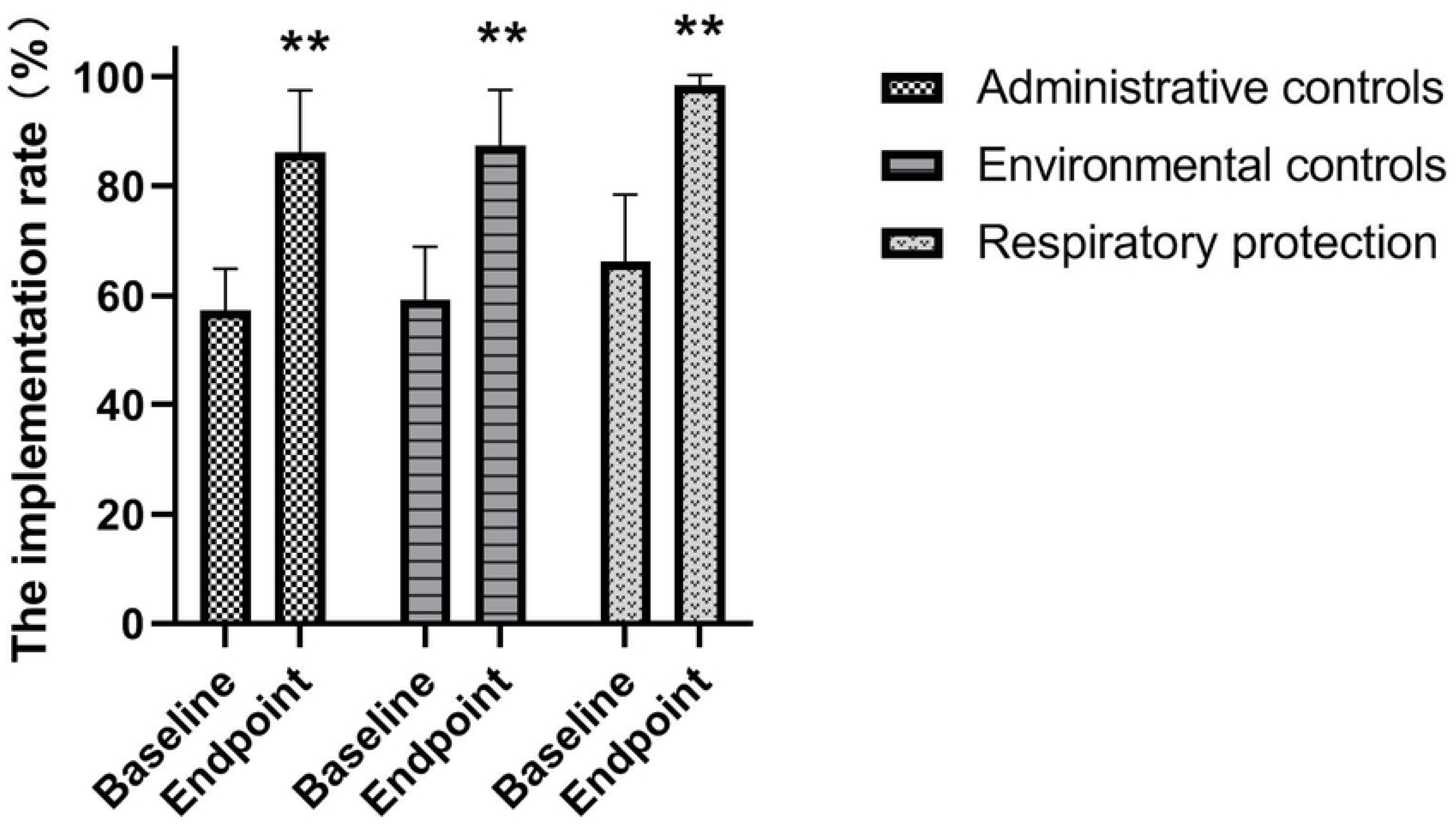
The improvement of TBIC in the designated hospitals. Data represents mean ± standard error of the mean. (**p < 0.01).

### 2. The improvement of TBIC in different hospitals

At baseline, the implementation rate of administrative controls, environmental controls as well as the use of respiratory protection were 54.34%, 69.59%, 73.58% in Suzhou Fifth People’s Hospital; 70.76%, 68.83%, 75.74% in Wuxi Fifth People’s Hospital; 48.19%, 44.92%, 51.29% in Changzhou Third People’s Hospital; 58.18%, 60.69%, 74.72% in Yangzhou Third People’s Hospital; 53.48%, 59.9%, 49.72% in Zhengjiang Third People’s Hospital; and 58.21%, 51.32%, 72.31% in Nantong Sixth People’s Hospital, respectively.

After six quarterly evaluations and implementation, the implementation rate of administrative controls, environmental controls as well as the use of respiratory protection were 70.73%, 84.37%, 98.14% in Suzhou Fifth People’s Hospital; 97.62%, 95.67%, 100% in Wuxi Fifth People’s Hospital; 77.41%, 68.05%, 95.46% in Changzhou Third People’s Hospital; 82.06%, 92.87%, 96.94% in Yangzhou Third People’s Hospital; 97.7%, 91.5%, 100% in Zhengjiang Third People’s Hospital; and 92.08%, 92%, 100% Nantong Sixth People’s Hospital, respectively (Fig 2).

**Fig 2.**
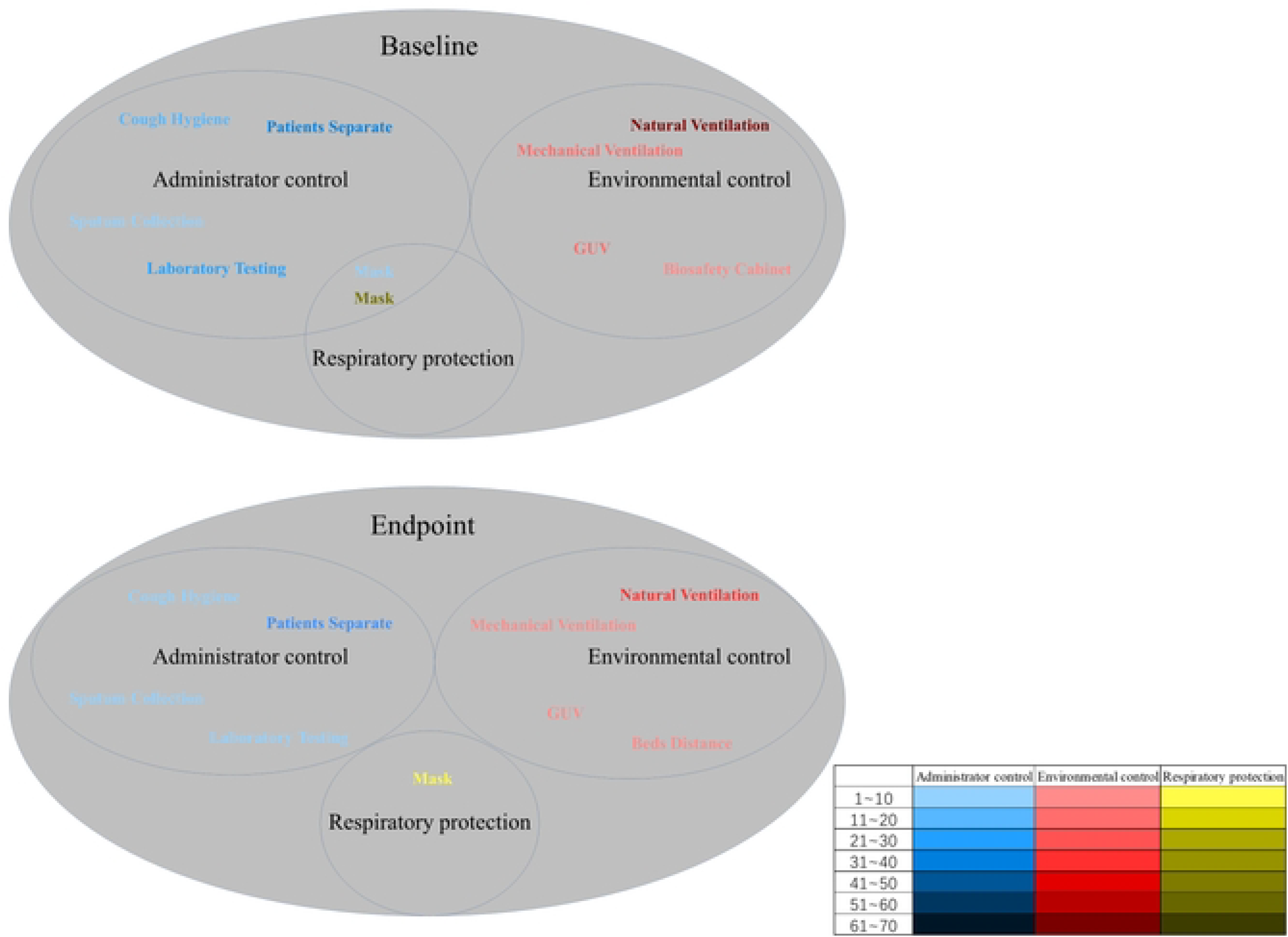
The improvement of TBIC in different hospitals

### 3. The improvement of TBIC in the departments of hospitals

At baseline, the implementation rate of administrative controls, environmental controls as well as the use of respiratory protection were 49.88%, 51.23%, 60.82% in TB outpatient department, 59.44%, 60.94%, 69.54% in inpatient department; and 71.25%, 65.42%, 68.33% in laboratory, respectively.

After six quarterly evaluation and implementation, the implementation rate of administrative controls, environmental controls as well as the use of respiratory protection were 83.54%, 81.9%, 99.31% in TB outpatient department; 85%, 92.98%, 100% in inpatient department; and 86.86%, 87.37%, 95.97% in laboratory, respectively (Fig 3).

**Fig 3.**
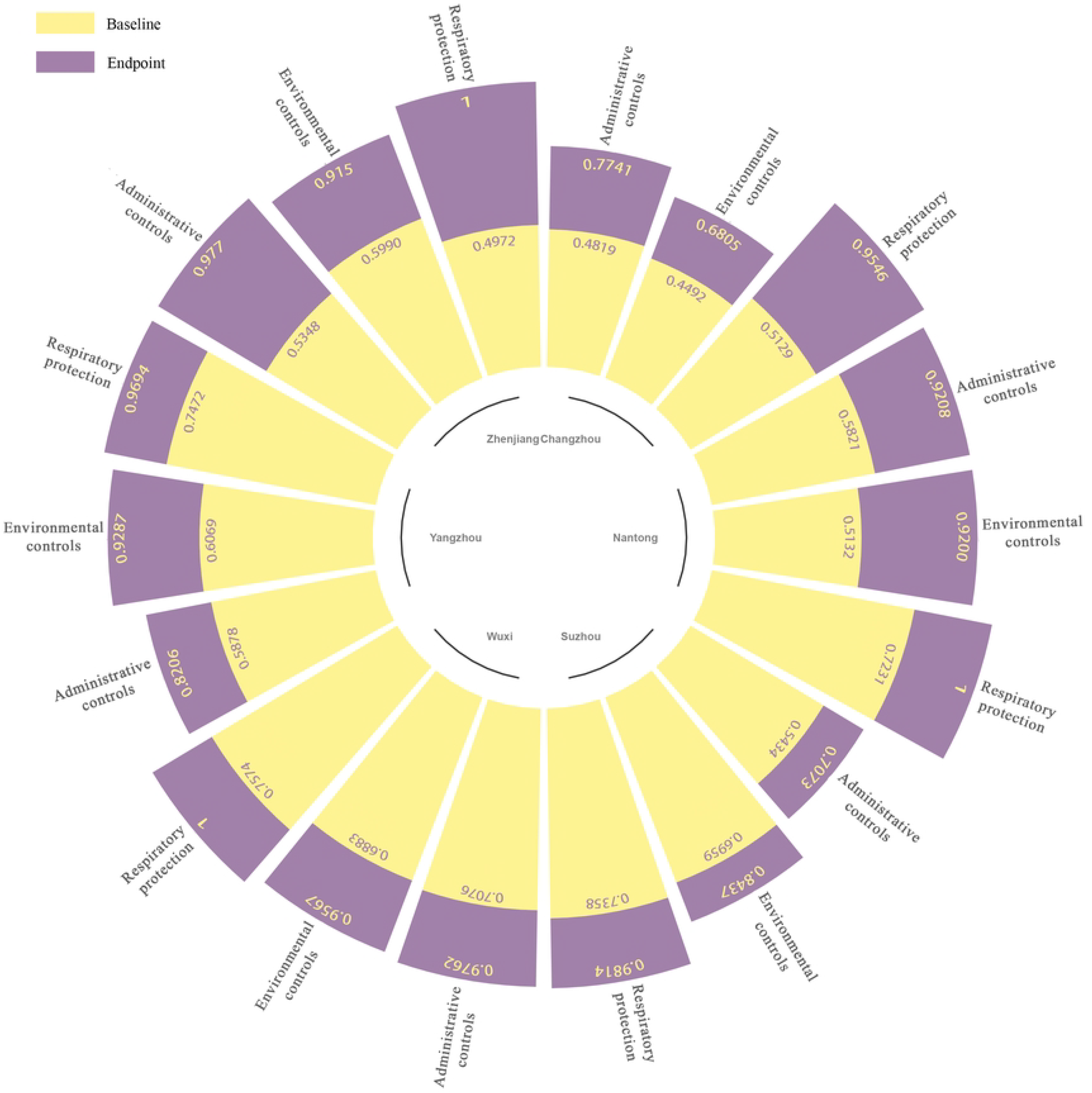
The improvement of TBIC in the departments of hospitals.

### 4. Analysis of main problems

We analyzed the main problems of different departments at the baseline and final evaluation, respectively. The main problems of administrator control include a lack of patients’ separation methods, a lack of cough hygiene education, and the feedback of laboratory test results takes a long time and the insufficient of sputum collection condition. The main problems of environment control include a lack of ventilation management (both natural and mechanical), a lack of UV lights in some areas, and a lack of maintenance of biosafety cabinets. The main problem of respiratory protection was a lack of respirator training. After 18 months’ evaluation and implementation, although some problems still exist, there was significant improvement comparing to the baseline assessment: priority way for cough patients was established, mechanical ventilation and the use of masks had been improved, the UV lights were settled in need, the UVGI was settled in the TB consulting room (Fig 4).

**Fig 4.**
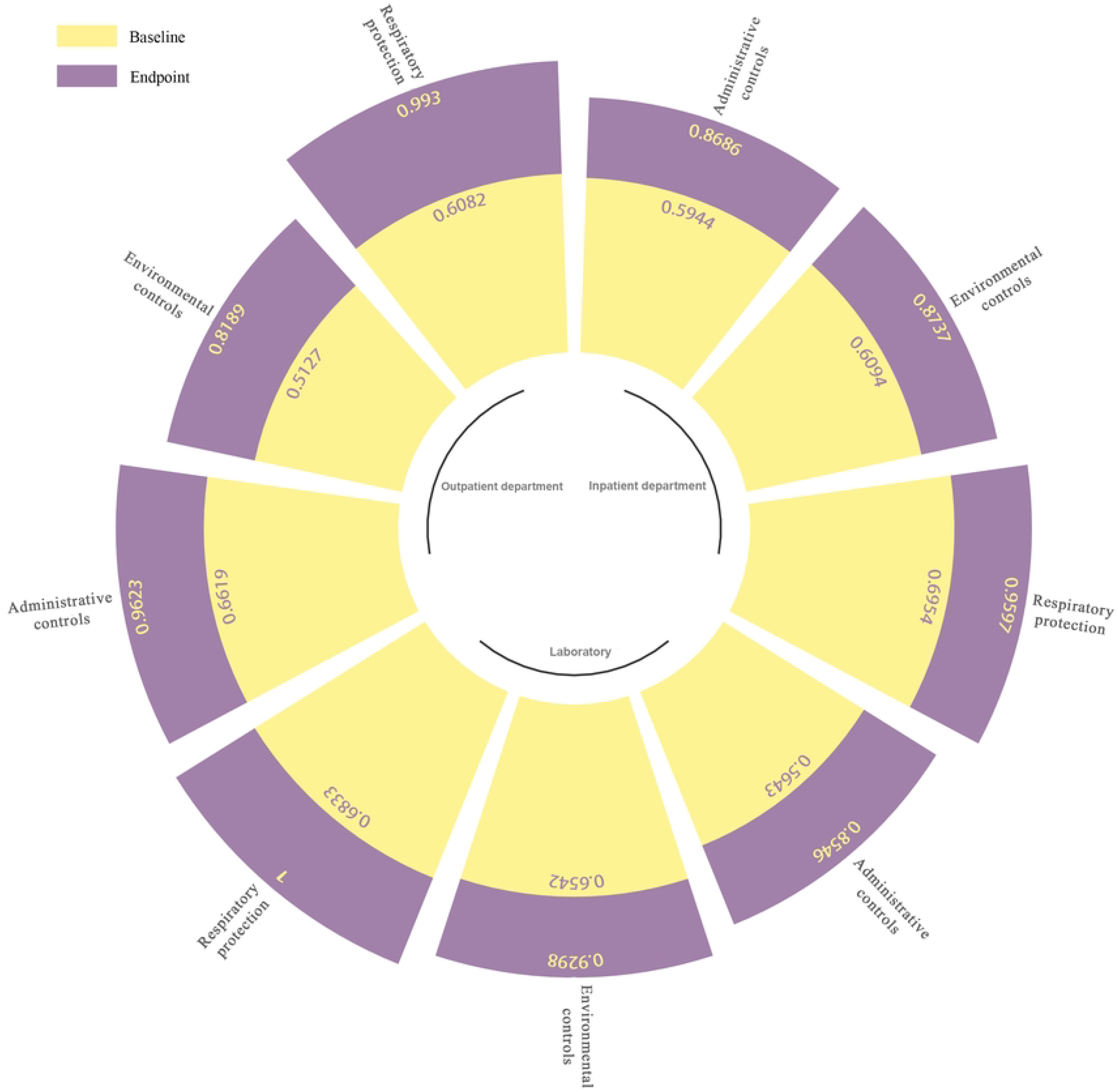
Analysis of the main problems at baseline and endpoint evaluation

## Discussion

TB infection control is a key element in the strategy to end the global TB epidemic[6], but there are still critical infection control gaps in health facilities, posing a continuous risk to personnels living with HIV infection, health care workers, and the uninfected[7-9]. Widespread implementation of infection control measures, especially in settings with high numbers of cases, should help prevent further TB transmission and ultimately bring the global TB epidemic to an end[10, 11].

In China, as in many countries with a high incidence of TB and limited public health resources, implementation of TB infection control measures has been inadequate[12, 13]. In addition, the TB designated hospital is also the COVID-19 designated hospital, since the outbreak of the COVID-19 pandemic[14]. So, infection control is particularly important. The aim at improving administrator control, environmental control, and respiratory protection is to reduce nosocomial transmission in hospitals.

Due to the impact of the COVID-19 pandemic, the complete time is variable in different hospitals, and the evaluation continued for more than 18 months. As the result, administrator control measures mainly involved implementation of existing policies, including no priority arrangement for cough patients, lack of isolation of TB inpatient department from other infectious diseases, lack of patients’ hygiene education, inadequate TBIC training for HCWs, our findings are similar to Paleckyte’s research [15]. The inadequacy was changed in practices and rapidly put into place. Environmental control measures mainly involved insufficient natural and mechanical ventilation in some consulting rooms and TB wards (<6 ACH) the shortage of UV lights; respiratory protection measures mainly involved appropriate PPE for some coughing patients as well as HCWs not carrying out fit tests of medical respirators, improvements were instituted at a minimal cost. After more than 18 months’ evaluation and implementation, a priority way for cough patients was established, ventilation and the use of masks had been improved, the UV lights were settled in need, the UVGI was settled in the TB consulting room and some TB wards. The implementation rate of administrative, environmental and PPE was significantly improved in all of the hospitals. The main problem was the inadequate of separate TB inpatient department, and the availability of separate TB wards remains suboptimal.

## Conclusions

The findings in this project demonstrated there are still many inadequacies of TBIC in designated hospitals, TB IC measures must be strengthened to reduce TB transmission among HCWs and non-TB patients. This method was practicable and suitable to be popularized in countries with a high burden of TB. The long-term impact and sustainability of the TB infection control practices implemented should be assessed.

## Data Availability

This text is appropriate if the data are owned by a third party and authors do not have permission to share the data. https://pan.baidu.com/s/1u38ik9oFepKQR8Gv2_Sjnw?pwd=gqol

## Funding

This work was supported by Strengthening TBIC Management and Special fund for tuberculosis in Jiangsu Province. Jiangsu Young Medical Talents Program (grant number: QNRC2016541) and Jiangsu Provincial Medical Key Discipline (ZDXK202250)

## Ethics Statements

This research is an indicator study in hospitals, which does not involve human or experimental animals.

## Conflict of Interest

No conflict of Interest

## Acknowledgments

We would like to thank Dr. Zikai Feng for his editorial assistant.

## Notes

### Competing Interest Statement

The authors have declared no competing interest.

### Funding Statement

Yes

